# Heart murmur detection from phonocardiogram recordings: The George B. Moody PhysioNet Challenge 2022

**DOI:** 10.1101/2022.08.11.22278688

**Authors:** Matthew A. Reyna, Yashar Kiarashi, Andoni Elola, Jorge Oliveira, Francesco Renna, Annie Gu, Erick A. Perez Alday, Nadi Sadr, Ashish Sharma, Jacques Kpodonu, Sandra Mattos, Miguel T. Coimbra, Reza Sameni, Ali Bahrami Rad, Gari D. Clifford

**Affiliations:** Department of Biomedical Informatics, Emory University, Atlanta, GA, USA; Department of Electronic Technology, University of the Basque Country UPV/EHU, Eibar, Gipuzkoa, Spain; REMIT, Universidade Portucalense, Porto, Portugal; INESC TEC, Faculdade de Ciências, Universidade do Porto, Porto, Portugal; ResMed, Sydney, Australia; Division of Cardiac Surgery, Beth Israel Deaconess Medical Center, Harvard Medical School, Boston, MA, USA; Unidade de Cardiologia e Medicina Fetal, Real Hospital Português, Recife, Pernambuco, Brazil; Department of Biomedical Engineering, Emory University and the Georgia Institute of Technology, Atlanta, GA, USA

**Author notes:** Department of Biomedical Informatics, Emory University, Atlanta, Georgia, United States. Deceased (Ashish Sharma).

## Abstract

Cardiac auscultation is an accessible diagnostic screening tool that can help to identify patients with heart murmurs for follow-up diagnostic screening and treatment for abnormal cardiac function. However, experts are needed to interpret the heart sounds, limiting the accessibility of auscultation for cardiac care in resource-constrained environments. Therefore, the George B. Moody PhysioNet Challenge 2022 invited teams to develop algorithmic approaches for detecting heart murmurs and abnormal cardiac function from phonocardiogram (PCG) recordings of the heart sounds.

For the Challenge, we sourced 5272 PCG recordings from 1568 pediatric patients in rural Brazil, and we invited teams to implement diagnostic screening algorithms for detecting heart murmurs and abnormal cardiac function from the recordings. We required the participants to submit the complete code for training and running their algorithms, improving the transparency, reproducibility, and utility of their work. We also devised an evaluation metric that considered the costs of screening, diagnosis, treatment, and diagnostic errors, allowing us to investigate the benefits of algorithmic diagnostic screening and facilitate the development of more clinically relevant algorithms.

We received 779 algorithms from 87 teams during the course of the Challenge, resulting in 53 working codebases for detecting heart murmurs and abnormal cardiac function from PCGs. These algorithms represent a diversity of approaches from both academia and industry.

The use of heart sound recordings for identifying heart murmurs and abnormal cardiac function allowed us to explore the potential of algorithmic approaches for providing accessible pre-screening in resource-constrained environments. The submission of working, open-source algorithms and the use of novel evaluation metrics supported the reproducibility, generalizability, and clinical relevance of the research from the Challenge.

**Author summary:** Cardiac auscultation is an accessible diagnostic screening tool for identifying heart murmurs. However, experts are needed to interpret heart sounds, limiting the accessibility of auscultation in cardiac care. The George B. Moody PhysioNet Challenge 2022 invited teams to develop algorithms for detecting heart murmurs and abnormal cardiac function from phonocardiogram (PCG) recordings of heart sounds.

For the Challenge, we sourced 5272 PCG recordings from 1568 pediatric patients in rural Brazil. We required the participants to submit the complete code for training and running their algorithms, improving the transparency, reproducibility, and utility of their work. We also devised an evaluation metric that considered the costs of screening, diagnosis, treatment, and diagnostic errors, allowing us to investigate the benefits of algorithmic diagnostic screening and facilitate the development of more clinically relevant algorithms. We received 779 algorithms from 87 teams during the Challenge, resulting in 53 working codebases and publications that represented a diversity of approaches to detecting heart murmurs and identifying clinical outcomes from heart sound recordings.

## Introduction

Cardiac auscultation via stethoscopes remains the most common and the most cost-effective tool for cardiac pre-screening. Despite its popularity, the technology has limited diagnostic sensitivity and accuracy [1, 2], as its interpretation requires many years of experience and skill, leading to significant disagreement between medical personnel [3, 4]. Digital phonocardiography has emerged as a more objective alternative to traditional cardiac auscultation, enabling the use of algorithmic methods for heart sound analysis and diagnosis [5]. The phonocardiogram (PCG) can be acquired by a combination of high-fidelity stethoscope front-ends and high-resolution digital sampling circuitry, which enable the registration of heart sounds as a discrete-time signal.

As acoustic signals, heart sounds are mainly generated by the vibrations of cardiac valves as they open and close during the cardiac cycle and by the turbulence of blood flow within the valves. Turbulent blood flow may cause enough vibrations within the cardiac valves to create audible heart sounds and abnormal waveforms in the PCG, which are called *murmurs*. Different kinds of murmurs exist, and they are characterized in various ways, including location, timing, duration, shape, intensity, and pitch. The identification and analysis of murmurs provide valuable information about cardiovascular pathologies.

However, while cardiac auscultation itself is relatively accessible, experts are needed to interpret heart sounds and PCGs, limiting the accessibility of auscultation for cardiac disease management, especially in resource-constrained environments. The ability to correctly interpret PCGs for murmur detection and for identifying different pathologies requires time and broad clinical experience. Therefore, the objective interpretation of the PCG remains a difficult skill to acquire.

The 2022 George B. Moody PhysioNet Challenge (formerly the PhysioNet/Computing in Cardiology Challenge) provided an opportunity to address these issues by inviting teams to develop fully automated approaches for detecting heart murmurs and abnormal cardiac function from PCG recordings. We sourced and shared PCG recordings for up to four auscultation locations from a largely pediatric population in Brazil, and we asked teams to identify both heart murmurs and the clinical outcomes of a full diagnostic screening from the recordings. The Challenge explored the diagnostic potential of algorithmic approaches for interpreting PCG recordings.

## Methods

### Challenge Data

The CirCor DigiScope dataset [6] was used for the 2022 George B. Moody PhysioNet Challenge. This dataset consists of one or more PCG recordings from several auscultation locations. The dataset was collected during two screening campaigns in Paraíba, Brazil from July 2014 to August 2014 and from June 2015 to July 2015. The study protocol was approved by the 5192-Complexo Hospitalar HUOC/PROCAPE Institutional Review Board under the request of the Real Hospital Português de Beneficiência em Pernambuco. A detailed description of the dataset can be found in [6].

During the data collection sessions, the participants answered a socio-demographic questionnaire, followed by a clinical examination (anamnesis and physical examination), a nursing assessment (physiological measurements), and cardiac investigations (cardiac auscultation, chest radiography, electrocardiogram, and echocardiogram). The collected data were then analyzed by an expert pediatric cardiologist. The expert could re-auscultate the participant or request complementary tests. At the end of the session, the pediatric cardiologist either directed the participant for a follow-up appointment, referred the participant to cardiac catheterization or heart surgery, or discharged the participant as appropriate.

The PCGs were recorded using an electronic auscultation device, the Littmann 3200 stethoscope, from up to four auscultation locations on the body; see Fig 1:

**Fig 1.**
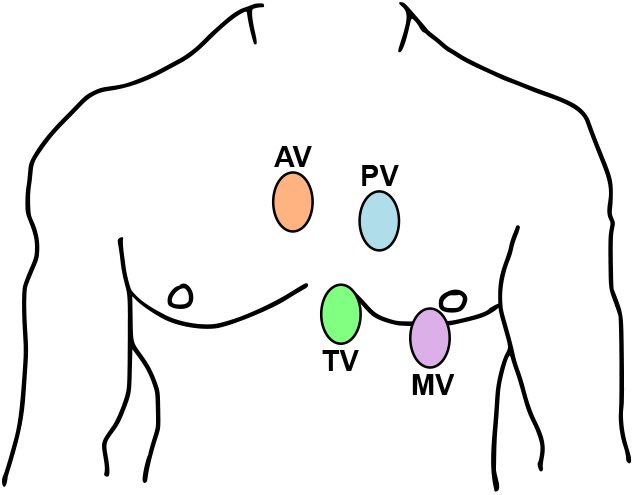
Auscultation locations for the CirCor DigiScope dataset [6], which was used for the Challenge: pulmonary valve (PV), aortic valve (AV), mitral valve (MV), and tricuspid valve (TV).

- Aortic valve: second intercostal space, right sternal border;
- Pulmonic valve: second intercostal space, left sternal border;
- Tricuspid valve: left lower sternal border; and
- Mitral valve: fifth intercostal space, midclavicular line (cardiac apex).

The choice of locations, the number of recordings at each location, and the duration of the PCG recordings varied between patients. The recordings were made by potentially different operators, but each patient’s PCGs was recorded by a single operator in a sequential manner. The PCG were also inspected for signal quality and semi-automatically segmented using the three algorithms proposed in [7], [8], and [9] and then corrected, as necessary, by a cardiac physiologist.

Each patient’s PCGs and clinical notes were also annotated for murmurs and abnormal cardiac function. These annotations served as the labels for the Challenge.

The murmur annotations and characteristics (location, timing, shape, pitch, quality, and grade) were manually identified by a single cardiac physiologist independently of the available clinical notes and PCG segmentation. The cardiac physiologist annotated the PCGs by listening to the audio recordings and by visually inspecting the corresponding waveforms. The murmur annotations indicated whether the expert annotator could detect the presence or absence of a murmur in a patient from the PCG recordings or whether the annotator was unsure about the presence or absence of a murmur. The murmur annotations did not indicate whether a murmur was pathological or innocent.

The clinical outcome annotations were determined by cardiac physiologists using all available clinical notes, including the socio-demographic questionnaire, clinical examination, nursing assessment, and cardiac investigations. In particular, the experts used reports from a echocardiogram, which is a standard diagnostic tool for identifying abnormal cardiac function. The clinical outcome annotations indicated whether the expert annotator identified normal or abnormal cardiac function. The clinical outcome annotations were performed by different experts, and these experts were different from the expert who performed the murmur annotations.

In total, the Challenge dataset consisted of 5272 annotated PCG recordings from 1568 patients. We released 60% of the recordings as a public training set and retained 10% of the recordings as a private validation set and 30% of the recordings as a private test set. The training, validation, and test sets were matched to approximately preserve the univariate distributions of the variables. Data from patients who participated in multiple screening campaigns belonged to only one of the training, validation, or test sets to prevent data leakage. We shared the training set at the beginning of the Challenge to allow the participants to develop their algorithms and sequestered the validation and test sets during the Challenge to evaluate the algorithms.

Table 1 summarizes the variables provided in the training, validation, and test sets of the Challenge data. Table 2 summarizes the distributions of the variables.

**Table 1.**
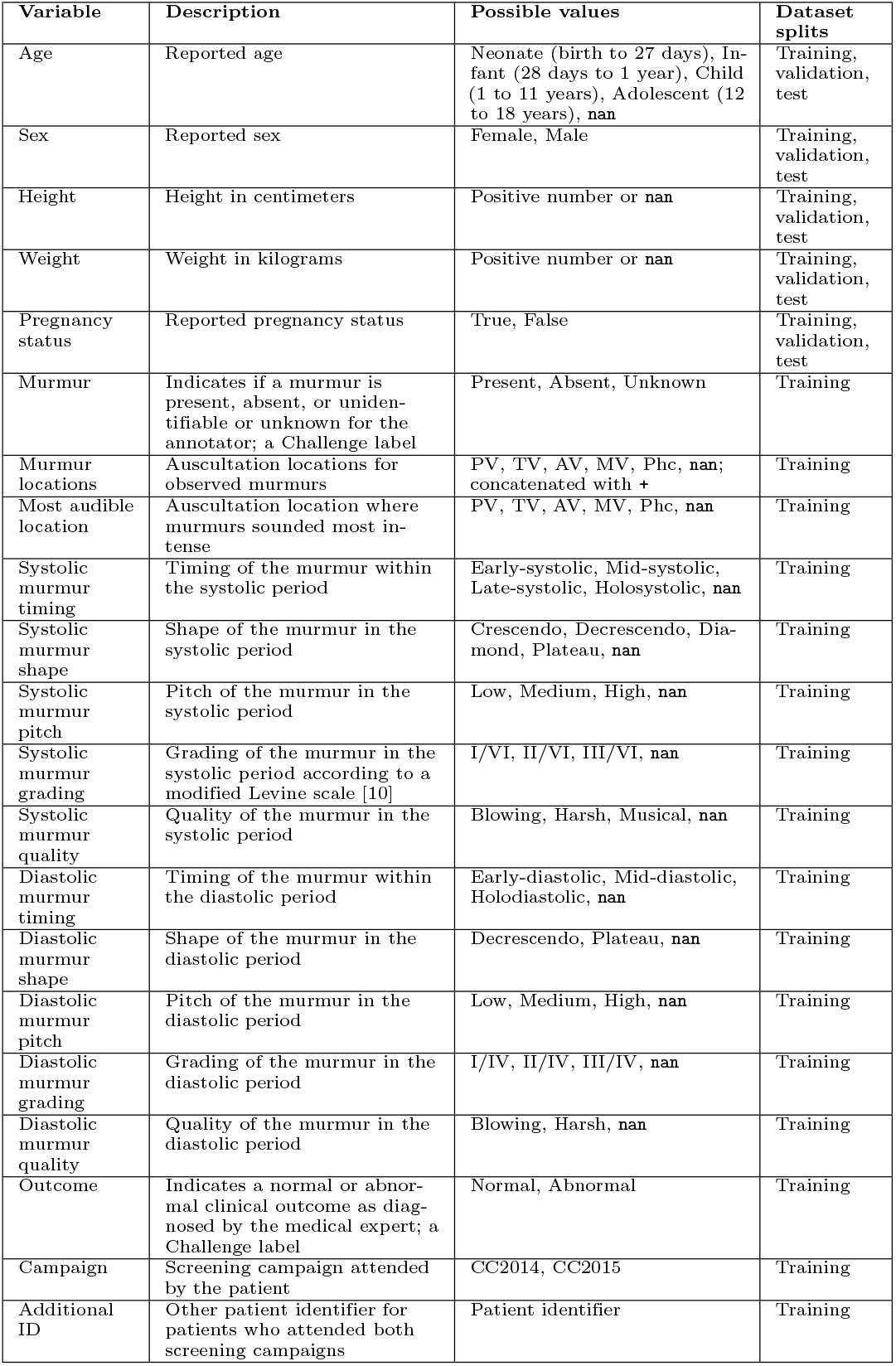
Demographic, murmur, and clinical outcome information in the Challenge training, validation, and/or test sets; `nan` values indicate unknown or missing values.

**Table 2.** Demographic, murmur, and clinical outcome distributions across the Challenge training, validation, and/or test data. For categorical variables, the entries of the table denote the fraction of the dataset with each possible value. For numerical variables, the entries of the table denote the median and first and third quartiles, respectively, of the values in the dataset, i.e., median [Q1, Q3].

| Variable | Training set | Validation set | Test set | Entire dataset |
| --- | --- | --- | --- | --- |
| Age |  |  |  |  |
| - Neonate | 0.006 | 0.007 | 0.006 | 0.006 |
| - Infant | 0.134 | 0.101 | 0.107 | 0.122 |
| - Child | 0.705 | 0.678 | 0.723 | 0.708 |
| - Adolescent | 0.076 | 0.094 | 0.099 | 0.085 |
| - nan | 0.079 | 0.121 | 0.065 | 0.078 |
| Sex |  |  |  |  |
| - Female | 0.516 | 0.530 | 0.453 | 0.498 |
| - Male | 0.484 | 0.470 | 0.547 | 0.502 |
| Height | 115 [89, 133] cm | 116 [89, 136] cm | 113 [92, 135] cm | 115 [89, 134] cm |
| Weight | 20.4 [12.5, 31.2] kg | 20.9 [12.9, 34.6] kg | 21.0 [13.4, 32.7] kg | 20.6 [12.7, 32.0] kg |
| Pregnancy status |  |  |  |  |
| - False | 0.926 | 0.899 | 0.948 | 0.930 |
| - True | 0.074 | 0.101 | 0.052 | 0.07 |
| Murmur |  |  |  |  |
| - Absent | 0.738 | 0.711 | 0.719 | 0.730 |
| - Unknown | 0.072 | 0.107 | 0.073 | 0.076 |
| - Present | 0.190 | 0.181 | 0.208 | 0.195 |
| Outcome |  |  |  |  |
| - Normal | 0.516 | 0.617 | 0.493 | 0.518 |
| - Abnormal | 0.484 | 0.383 | 0.507 | 0.482 |
| Campaign |  |  |  |  |
| - CC2014 | 0.409 | 0.403 | 0.436 | 0.416 |
| - CC2015 | 0.591 | 0.597 | 0.564 | 0.584 |

### Challenge Objective

We designed the Challenge to explore the potential for algorithmic pre-screening of heart murmurs and abnormal heart function, especially in resource-constrained environments. We asked the Challenge participants to design working, open-source algorithms for identifying heart murmurs and the clinical outcomes from PCG recordings. For each patient encounter, each algorithm interpreted the PCG recordings and/or demographic data.

#### Challenge Timeline

This year’s Challenge was the 23^rd^ George B. Moody PhysioNet Challenge [11]. As with previous years, this year’s Challenge had an unofficial phase and an official phase.

The unofficial phase (February 1, 2022 to April 8, 2022) introduced the teams to the Challenge. We publicly shared the Challenge objective, training data, example classifiers, and evaluation metrics at the beginning of the unofficial phase. We invited the teams to submit their code for evaluation, and we scored at most 5 entries from each team on the hidden validation set during the unofficial phase.

Between the unofficial phase and official phase, we took a hiatus (April 9, 2022 to April 30, 2022) to improve the Challenge in response to feedback from teams, the broader community, and our collaborators. During the hiatus, we sourced the clinical outcomes for the patient encounters to enrich the Challenge.

The official phase (May 1, 2022 to August 15, 2022) allowed the teams to refine their approaches for the Challenge. We updated the Challenge objectives, data, example classifiers, and evaluation metric at the beginning of the official phase. We again invited the teams to submit their code for evaluation, and we scored at most 10 entries from each team on the hidden validation set during the official phase.

After the end of the official phase, we asked each team to choose a single entry from their team for evaluation on the test set. We allowed the teams to choose any successful model from the official phase, but most teams chose their best-scoring entries. We only evaluated one entry from each team for each task on the test set to prevent sequential training on the test set. The winners of the Challenge were the teams with the best scores on the test set. We announced the winners at the end of the Computing in Cardiology (CinC) 2022 conference.

The teams presented and defended their work at CinC 2022, and they wrote four-page conference proceeding papers describing their work, which we reviewed for accuracy and coherence. We will publicly release the algorithms after the end of the Challenge and the publication of these papers through the Challenge website.

#### Challenge Rules and Expectations

While we encouraged the teams to ask questions, pose concerns, and discuss the Challenge in a public forum, we prohibited the teams from discussing or sharing their work during the unofficial phase, hiatus, and official phase of the Challenge to preserve the diversity and uniqueness of the teams’ approaches.

For both phases of the Challenge, we required teams to submit their code for training and running their models, including any code for processing or relabeling the data. We first ran each team’s training code on the public training set to create trained models. We then ran the trained models on the hidden validation and test sets to label the recordings; we ran the trained models on the recordings sequentially to better reflect the sequential nature of the screening process. We then scored the outputs from the models using the expert annotations on hidden validation and test sets.

We allowed the teams to submit either MATLAB or Python code; other implementations were considered upon request, but there were no requests for other programming languages. Participants containerized their code in Docker and submitted it by sharing private GitHub or Gitlab repositories with their code. We downloaded their code and ran it in containerized environments on Google Cloud. We described the computational architecture of these environments entries more fully in [12].

Each entry had access to 8 virtual CPUs, 52 GB RAM, 50 GB local storage, and an optional NVIDIA T4 Tensor Core GPU (driver version 470.82.01) with 16 GB VRAM. We imposed a 72 hour time limit for training each model on the training set without a GPU, a 48 hour time limit for training each model on the training set with a GPU, and a 24 hour time limit for running each trained model on either the validation or test set either with or without a GPU.

To aid the teams, we shared example MATLAB and Python entries. These examples used random forest classifiers with the age group, sex, height, weight, pregnancy status of the patient as well as the presence, mean, variance, and skewness, i.e., the first four order statistics, of the numerical values in each PCG recording as features. We did not design these example entries to perform well. Instead, we used them to provide minimal working examples of how to read the Challenge data and write the model outputs.

#### Challenge Evaluation

To capture the focus of this year’s Challenge on algorithmic pre-screening, we developed novel scoring metrics for each of the two Challenge tasks: detecting heart murmurs and identifying clinical outcomes for abnormal or normal heart function.

As described above, the murmurs were directly observable from the PCGs, but the clinical outcomes reflected the results of a more comprehensive diagnostic screening, including the interpretation of an echocardiogram. However, despite these differences, we asked the teams to perform both tasks using only PCGs and routine demographic data, which allowed us to explore the diagnostic potential of algorithmic approaches for interpreting relatively easily accessible PCGs.

The algorithms for both of these tasks effectively pre-screened patients for expert referral. If an algorithm inferred abnormal cardiac function, i.e., the murmur outputs were murmur present, murmur unknown, or outcome abnormal, then the algorithm would refer the patient to a human expert for a confirmatory diagnosis and potential treatment. If the algorithm inferred normal cardiac function, i.e., if the model outputs were murmur absent or outcome normal, then the algorithm would not refer the patient to an expert, and the patient would not receive treatment, even if the patient actually had abnormal cardiac function. Fig. 2 illustrates this algorithmic pre-screening process as part of a larger diagnostic pipeline.

**Fig 2.**
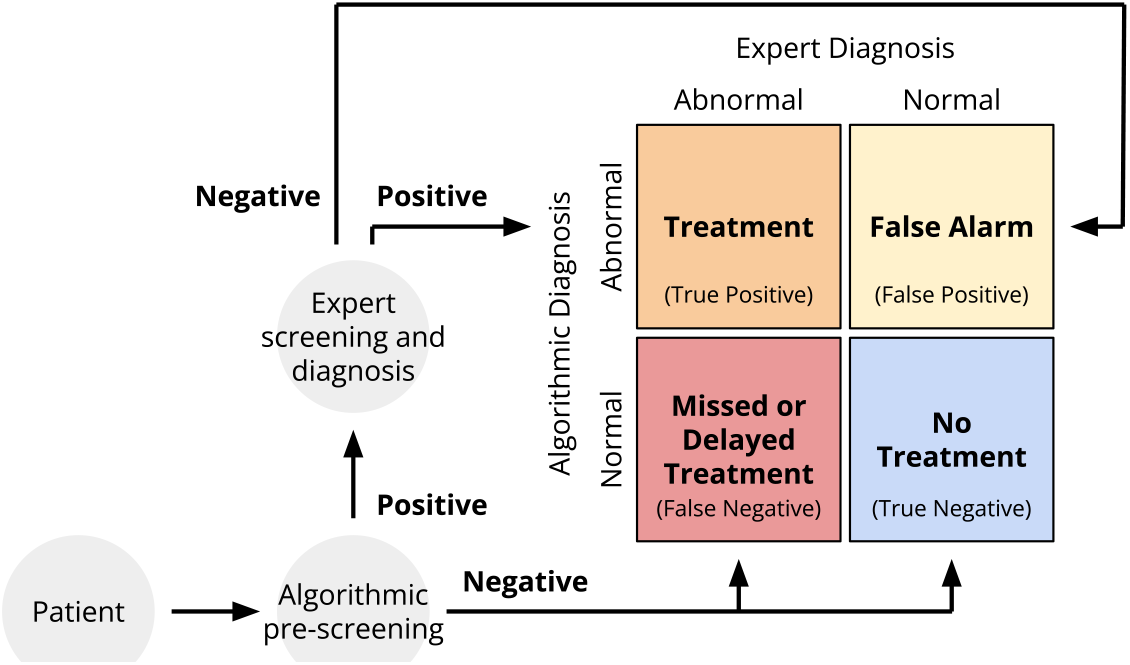
Screening and diagnosis pipeline for the Challenge. All patients would receive algorithmic pre-screening, and patients with positive results from algorithmic pre-screening would receive confirmatory expert screening and diagnosis. (i) Patients with positive results from algorithmic pre-screening and expert annotators would receive treatment; they are true positive cases. Patients with positive results from algorithmic pre-screening and negative results from expert annotators would not receive treatment; they are false positive cases or false alarms. Patients with negative results from algorithmic pre-screening who would have received positive results from the expert annotators would have missed or delayed treatment; they are false negative cases.

For the murmur detection task, we introduced a weighted accuracy metric that assessed the ability of an algorithm to reproduce the results of a skilled human annotator. For the clinical outcome identification task, we introduced a cost-based scoring metric that reflected the cost of expert diagnostic screening as well as the costs of timely, delayed, and missed treatments for abnormal cardiac function. The team with the highest weighted accuracy metric won the murmur detection task, and the team with the lowest cost-based scoring metric won the clinical outcome identification task.

We formulated versions of both of these metrics for both tasks to allow for more direct comparisons; see the Appendix for the additional metrics. We also calculated several traditional evaluation metrics to provide additional context to the performance of the models

Cost-based scoring is controversial, in part, because healthcare costs are an imperfect proxy for health needs [13, 14]; we reflect on this important issue in the Section. However, screening costs necessarily limit the ability to perform screening, especially in resource-constrained environments, so we considered costs as part of improving access to cardiac screening.

##### Weighted Accuracy Metric

We introduced a weighted accuracy metric to evaluate the murmur detection algorithms. This metric assessed the ability of the algorithms to reproduce the decisions of the expert annotator. This weighted accuracy metric is similar to the traditional accuracy metric, but it assigned more weight to patients that had or potentially had murmurs than to patients that did not have murmurs. These weights reflect the rationale that, in general, a missed diagnosis is more harmful than a false alarm.

Patients with negative results from algorithmic pre-screening who would have also received negative results from expert annotators also would not receive treatment; they are true negative cases.

We defined a weighted accuracy metric for the murmur detection task as

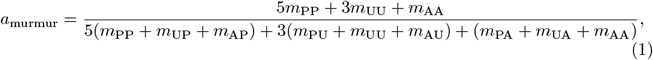

where Table 3 defines a three-by-three confusion matrix *M* = [*m*_*ij*_] for the murmur present, murmur unknown, and murmur absent classes.

**Table 3.** Confusion matrix *M* for murmur detection with three classes: murmur present, murmur unknown, and murmur absent. The columns are the ground truth labels from the human annotator, and the rows are the model outputs. The entries of the confusion matrix provide the number of patients with each model output for each ground truth label.

|  |  | Murmur Expert |  |  |
| --- | --- | --- | --- | --- |
|  |  | Present | Unknown | Absent |
| Murmur Classifier | Present | $m_{PP}$ | $m_{PU}$ | $m_{PA}$ |
| | Unknown | $m_{UP}$ | $m_{UU}$ | $m_{UA}$ |
| | Absent | $m_{AP}$ | $m_{AU}$ | $m_{AA}$ |

The coefficients were chosen to reflect the trade-off between false positives and false negatives, where clinicians may tolerate multiple false alarms to avoid a single missed diagnosis. In (1), murmur present cases have five times the weight of murmur absent cases (and the murmur unknown cases have three times the weight of murmur absent cases) to reflect a tolerance of five false alarms for every one false positive.

Like the traditional accuracy metric, this metric only rewarded algorithms for correctly classified recordings, but it provided the highest reward for correctly classifying recordings with murmurs and the lowest reward for correctly classifying recordings without murmurs, i.e., recordings that were labeled as having or not having murmurs, respectively. It provided an intermediate reward for correctly classifying recordings of unknown murmur status to reflect the difficulty and importance of indicating when the quality of a recording is not adequate for diagnosis.

We used (1) to rank the Challenge algorithms for the murmur detection task. The team with the highest value of (1) won this task.

##### Cost-based evaluation metric

We introduced a cost-based evaluation metric to evaluate the clinical outcome algorithms for abnormal or normal heart function. This metric considered the ability of the algorithms to reduce the costs associated with diagnosing and treating patients, primarily by screening fewer patients with normal cardiac function. We again emphasize that healthcare costs are an imperfect surrogate for health needs [13, 14]. However, costs are still a necessary consideration as part of resource-constrained environments.

For each patient encounter, the algorithm interpreted the PCG recordings and demographic data for the encounter. If an algorithm inferred abnormal cardiac function, then it would refer the patient to a human expert for a confirmatory diagnosis. If the expert confirmed the diagnosis, then the patient would receive treatment, and if the expert did not confirm the diagnosis, then the patient would not receive treatment.

Alternatively, if the algorithm inferred normal cardiac function, then it would not refer the patient to an expert, and the patient would not receive treatment, even if the patient had abnormal cardiac function that would have been detected by a human expert.

We associated each of these steps with a cost: the cost of algorithmic pre-screening, the cost of expert screening, the cost of timely treatment, and the cost of delayed or missed treatment.

For simplicity, we assumed that algorithmic pre-screening had a relatively small cost that depended linearly on the number of algorithmic pre-screenings. We also assumed that both timely treatments and delayed or missed treatments had relatively large costs that, on average, depended linearly on the number of individuals. Given our focus on the ability of algorithmic pre-screening to reduce human screening of patients with normal cardiac function, we assumed that expert screening had an intermediate cost that depended non-linearly on the number of screenings as well on as the infrastructure and capacity of the healthcare system. Of course, treatment costs are also non-linear in the number of treated patients for similar reasons, but non-urgent treatments can arguably utilize the capacity of the broader healthcare system. Screening far below the capacity of the healthcare system was inefficient and incurred a low total cost but high average cost. Screening above the capacity of the healthcare system was also inefficient and incurred both a high average cost and a high total cost.

Therefore, we introduced the following cost-based evaluation metric for identifying clinical outcomes. We defined the total cost of diagnosis and treatment with algorithmic pre-screening as

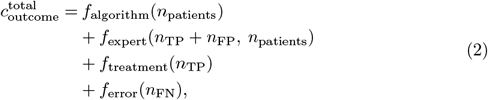

where Table 4 defines a two-by-two confusion matrix *N* = [*n*_*ij*_] for the clinical outcome abnormal and normal classes, *n*_patients_ = *n*_TP_ + *n*_FP_ + *n*_FN_ + *n*_TN_ is the total number of patients, and *f*_algorithm_, *f*_expert_, *f*_treatment_, *f*_error_ are defined below.

**Table 4.**
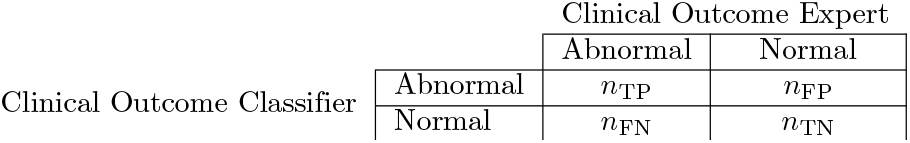
Confusion matrix *N* for clinical outcome detection with two classes: clinical outcome abnormal and clinical outcome normal. The columns are the ground truth labels from the human annotator, and the rows are the classifier outputs. The entries of the confusion matrix provide the number of patients with each classifier output for each ground truth label.

Again, for simplicity, we assumed that the costs for algorithmic pre-screening, timely treatments, and missed or late treatments were linear. We defined

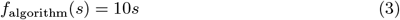

as the total cost of *s* pre-screenings by an algorithm,

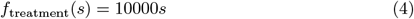

as the total cost of *s* treatments, and

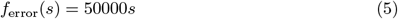

as the total cost of *s* missed or delayed treatments.

To better capture the potential benefits of algorithmic pre-screening, we assumed that the cost for expert screening was non-linear. We defined

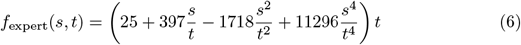

as the total cost of *s* screenings by a human expert out of a population of *t* patients so that

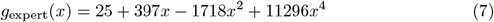

was the mean cost of screenings by a human expert when *x* = *s/t* of the patient cohort receives expert screenings; this reparameterization of (6) allowed us to compare algorithms on datasets with different numbers of patients. We designed (6) and (7) so that the mean cost of an expert screening was lowest when only 25% of the patient cohort received expert screenings but higher when screening below and above the capacity of the system. Fig 3 and Fig 4 show these costs across different patient cohort and screening sizes, and the Appendix provides a fuller derivation of (6) and (7).

**Fig 3.**
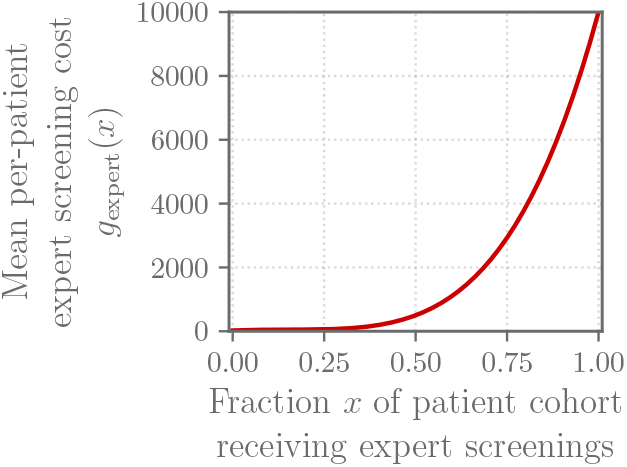
The expert screening cost *g*_expert_(*x*) defined for the Challenge: mean cost for screening a fraction *x* of a patient cohort for cardiac abnormalities. Mean per-patient expert screening cost *g*_expert_(*x*), i.e., the total expert screening cost for a patient cohort divided by the number of patients in the cohort.

**Fig 4.**
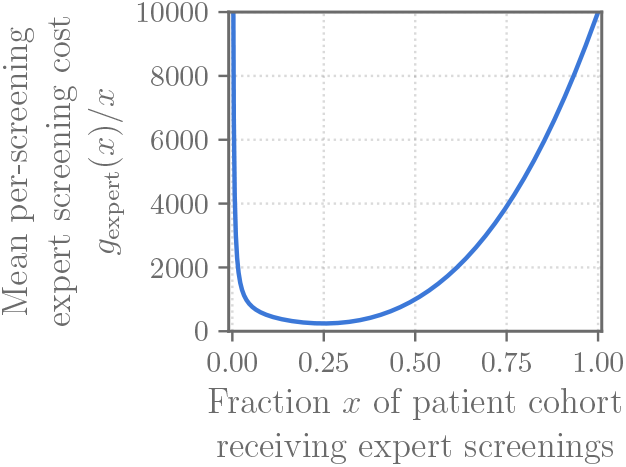
The expert screening cost *g*_expert_(*x*) defined for the Challenge: mean cost for screening a fraction *x* of a patient cohort for cardiac abnormalities. Mean per-screening expert screening cost *g*_expert_(*x*)*/x*, i.e., the total expert screening cost for a patient cohort divided by the number of patients in the cohort and the fraction of screenings in the cohort.

To compare costs for databases with different numbers of patients, e.g., the training, validation, and test databases, we defined the mean per-patient cost of diagnosis and treatment with algorithmic pre-screening as

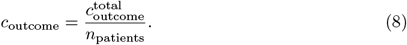

We used (8) to rank the Challenge algorithms for the murmur detection task. The team with the lowest value of (8) won this task.

### Challenge Results

#### Challenge Entries

We received 779 entries from 87 teams throughout the course of the 2022 PhysioNet Challenge, resulting in 77 submitted CinC abstracts, 62 accepted CinC abstracts, 43 published CinC proceedings papers, and 53 final codebases from 53 different teams.

These entries represent a diversity of approaches to the Challenge. A total of 81 teams submitted 293 entries during the unofficial phase of the Challenge, and a total of 63 teams submitted 486 entries during the official phase of the Challenge. Of the 779 entries, we received 652 entries from 75 teams that were implemented in Python, and 127 entries from 17 teams that were implemented in MATLAB. We received 519 entries from 60 teams that requested a graphics processing unit (GPU) for their entries, and 260 entries from 49 teams requested only a central processing unit (CPU), i.e., no GPU. In total, we received 473 successful entries that we were able to train on the public training set and evaluate on the hidden validation set and 306 entries that we were unable to train on the training set and/or evaluate on the validation validation set due to various errors in the submitted code.

A total of 58 teams had a successful entry during the official phase that we were able to train on the training set and evaluate on the validation set. Each team with a successful entry during the official phase selected a single entry for evaluation on the test set. Most teams chose their best-scoring entry from the official phase, but some teams selected their most recent entry or another entry because the entry with the best score on the validation set may not be the entry with the best score on the test set.

In some cases, a team’s best-scoring entry for the murmur detection task was different from the team’s best-scoring entry for the clinical outcome identification task because of the differences between the tasks and the evaluation metrics for grading them; in these cases, we chose a different entry for each task. However, since the teams could implement different approaches for each task, and since they could have submitted the different approaches as part of the same entry, we did not distinguish between teams and entries that submitted their best scoring entries in the same code submission or different code submissions. We will share the best scoring entries for both whether they were part of the same or different code submissions.

Of the 58 teams with a successful entry during the official phase, 53 teams had code that we were able to score on the training, validation, and test sets for both the murmur detection and outcome identification tasks, resulting in 53 working entries to the Challenge.

Fig 5 compares the performance of the working Challenge entries on the murmur detection and clinical outcome identification tasks. Fig 6 and Fig 7 compare the Challenge weighted accuracy and cost metrics with traditional scoring metrics, respectively, including area under the receiver operating characteristic curve (AUROC), area under the precision recall curve (AUPRC), the macro-averaged *F* -measure, and the traditional accuracy metric.

**Fig 5.**
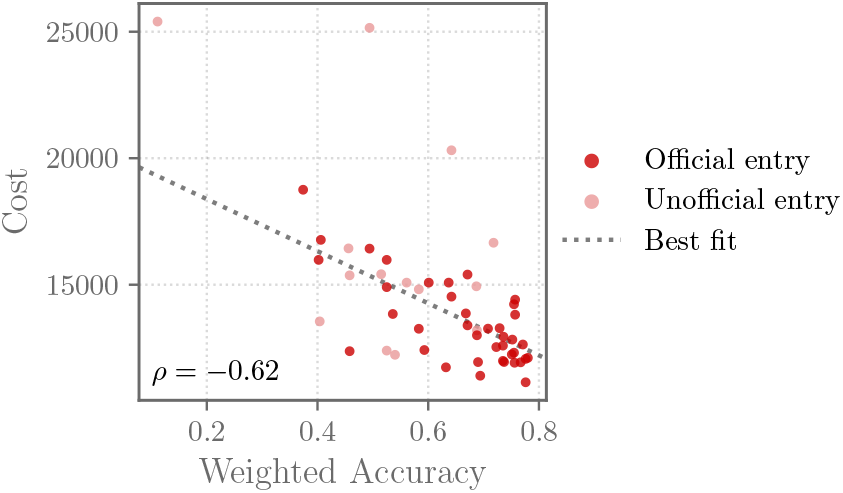
Weighted accuracy metric (1) for the murmur detection task (*x*-axis) and the cost metric (8) scores for the clinical outcome identification task (*y*-axis) of the final Challenge entries on the hidden test set. Each point shows an entry, and the shading of each point shows whether the entry was an official entry (dark red points) that satisfied all of the Challenge rules or an unofficial entry (light red points) that did not. The Spearman correlation coefficient *ρ* between the scores is given in the plot, and a line of best fit (gray dotted line) is given by a robust linear regression.

**Fig 6.**
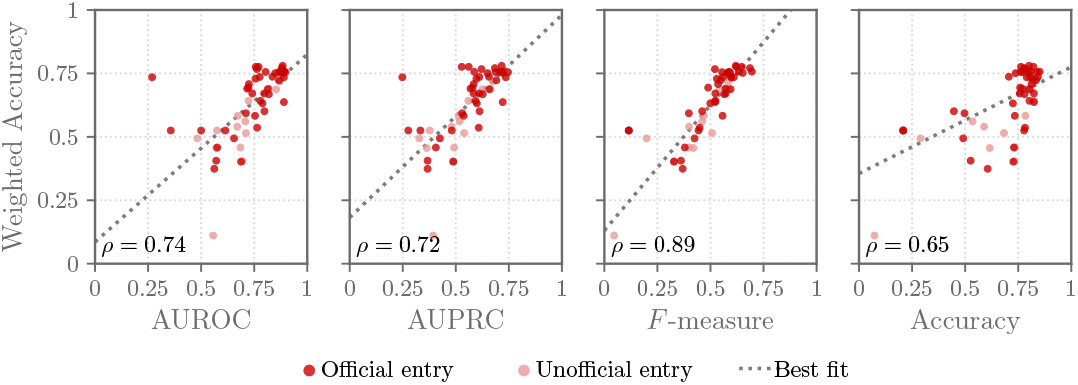
Traditional evaluation metric (*x*-axis) and weighted accuracy metric (*y*-axis) scores for the final Challenge entries with the murmur detection task on the hidden test set. AUROC is area under the receiving operating character curve, AUPRC is area under the precision-recall curve, *F* -measure is the macro-averaged *F* -measure, and accuracy is the traditional accuracy metric. Each point shows an entry, and the shading of each point shows whether the entry was an official entry (dark red points) that satisfied all of the Challenge rules or an unofficial entry (light red points) that did not. The Spearman correlation coefficients *ρ* between the metrics are given in the individual plots, and a line of best fit (gray dotted line) is given by a robust linear regression.

**Fig 7.**
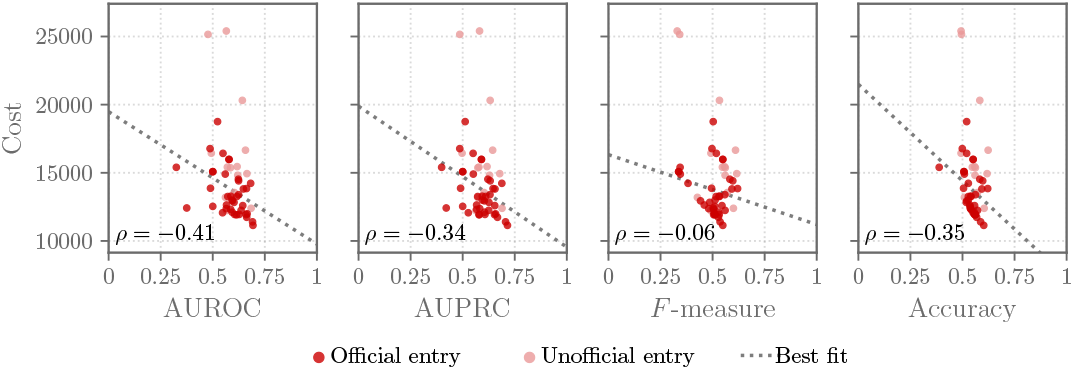
Traditional evaluation metric (*x*-axis) and cost metric (*y*-axis) scores for the final Challenge entries with the clinical outcome identification task on the hidden test set. AUROC is area under the receiving operating character curve, AUPRC is area under the precision-recall curve, *F* -measure is the traditional *F* -measure, and accuracy is the traditional accuracy metric. Each point shows an entry, and the shading of each point shows whether the entry was an official entry (dark red points) that satisfied all of the Challenge rules or an unofficial entry (light red points) that did not. The Spearman correlation coefficients *ρ* between the metrics are given in the individual plots, and a line of best fit (gray dotted line) is given by a robust linear regression.

To be officially ranked, we also required the teams to have successful unofficial and official phase entries, have an accepted CinC abstract, publicly share their CinC proceedings paper preprint by the CinC preprint submission deadline, have an accepted final CinC proceedings paper by the CinC final paper deadline, and license their code under an open source or similar license. We also allowed teams without a successful unofficial phase entry and/or accepted CinC abstract who had a successful, high-scoring official phase entry to submit a CinC abstract after the original abstract submission deadline as a “wild card” team. The abstract and proceedings paper was subject to the same review as the other participants.

We imposed these requirements in addition to the requirement for working code to support the dissemination of research into the research community. Of the 53 working entries, a total of 40 teams were officially ranked. Tables 5 and 6 summarize the traditional and Challenge evaluation metrics for the officially ranked entries for the murmur detection and clinical outcome identification tasks, respectively, on the hidden test set.

**Table 5.**
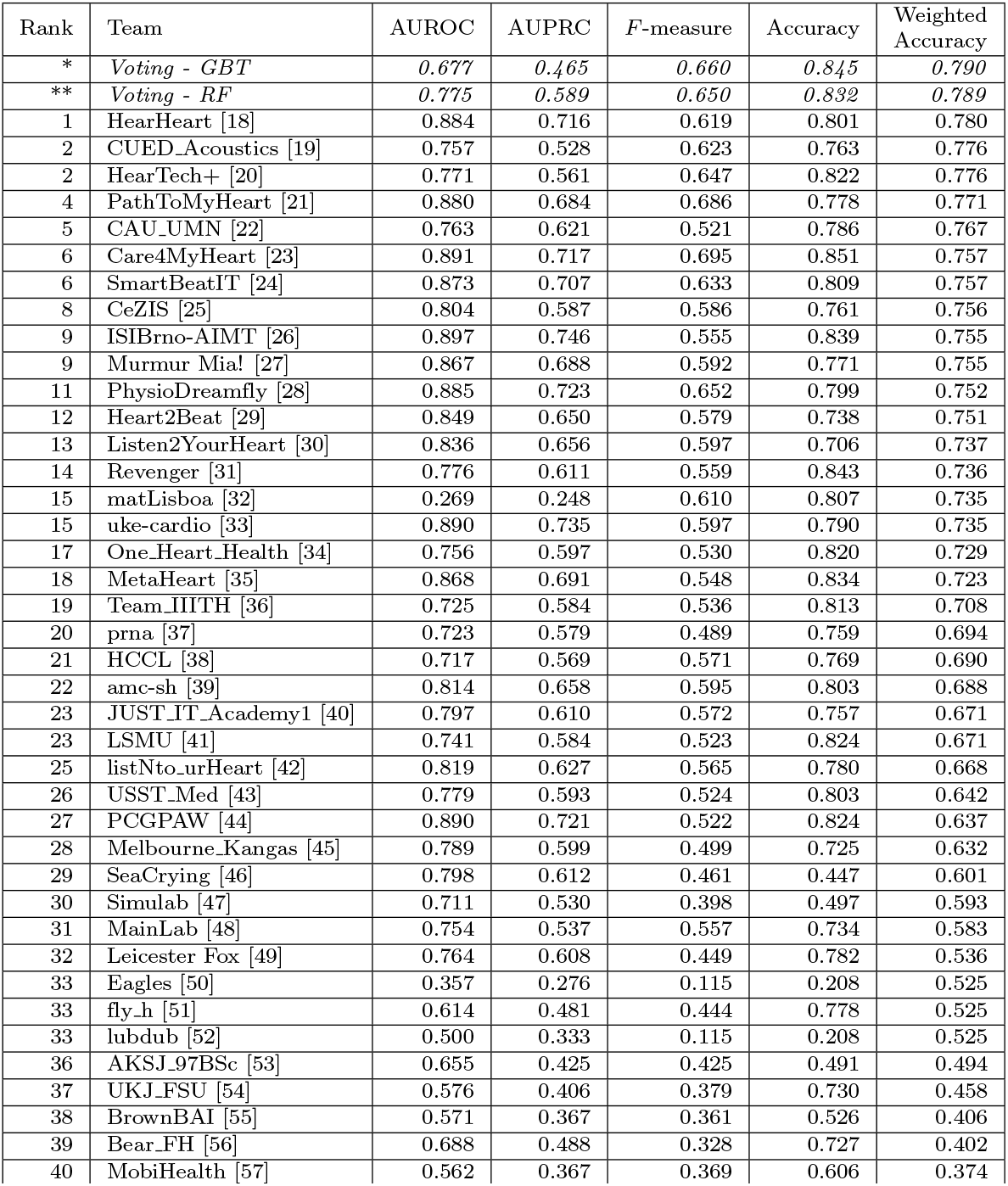
Scores of the officially ranked methods on the test set for the murmur detection task. AUROC is the area under the receiver operating characteristic curve, AUPRC is the area under the precision-recall curve, *F* -measure is the macro-averaged *F* -measure, accuracy is the traditional accuracy, and weighted accuracy is the weighted accuracy metric (1).

**Table 6.**
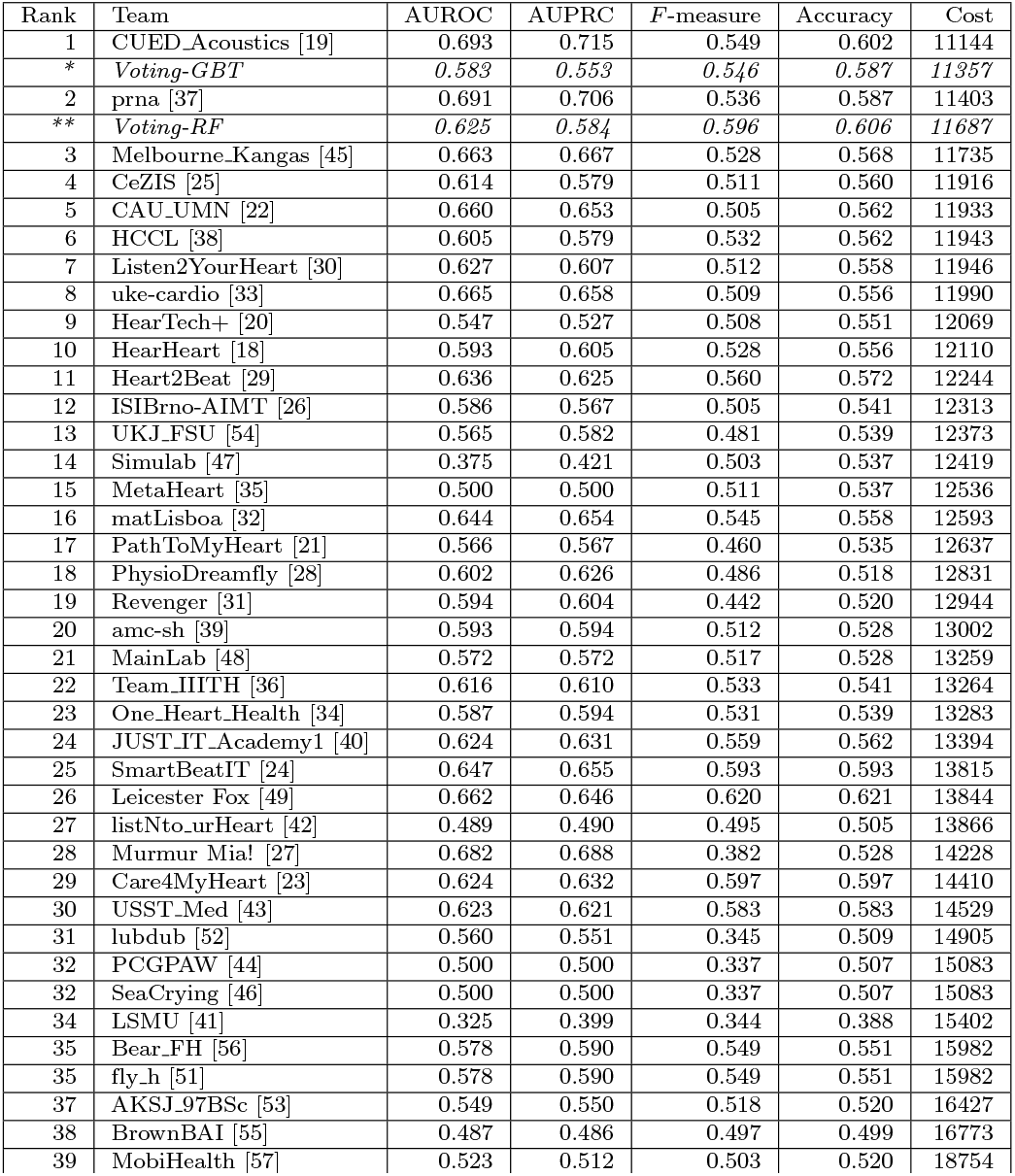
Scores of the officially ranked methods on the test set for the clinical outcome identification task. AUROC is the area under the receiver operating characteristic curve, AUPRC is the area under the precision-recall curve, *F* -measure is the traditional *F* -measure, accuracy is the traditional accuracy metric, and cost is the cost-based evaluation metric (8).

To assess the robustness of the algorithms, we also trained them on a subset of the training set with permuted labels and ran the retrained models on the validation and test sets with the original labels; we did not change any of the publicly available training data or labels but simply the size of the training set and the relationship between the data and the labels in the training set. Algorithms that truly learned from the training data performed much worse when retrained on the modified training set, while algorithms that did not learn from the training set performed the same or better. Only 35 of the 53 working entries were robust enough for us to complete this process. The remaining 18 working entries either crashed or achieving the same or a better score when retrained on the modified training set. While we encouraged teams to submit robust code, we did not inform teams *a priori* how we would test the robustness of their algorithms to avoid “gaming” the robustness criteria. Therefore, since we did not provide advance notice of the exact requirements for robustness, we did not disqualify teams whose algorithms failed this test.

#### Voting algorithms

While we ranked the official entries for the Challenge, we also recognized that entries with lower overall performance could achieve higher performance than the top-ranked entries on certain patient subgroups. Therefore, an algorithm that can learn the different strengths of different algorithms can improve on the overall performance of the individual entries. We developed several voting algorithms to leverage the diversity of Challenge entries.

For the voting algorithms, we considered the discrete classifier outputs (see Tables 3 and 4) from the 39 algorithms from teams that were officially ranked for both the murmur detection and clinical outcome tasks (see Tables 5 and 6). We considered these tasks separately, and we used the relevant Challenge scoring metric to evaluate the voting algorithms for each task: the weighted accuracy metric (1) for the murmur detection task and the cost metric (8) for the clinical outcome identification task. We used the common gradient-boosting trees (GBT) and random forests (RF) models for the voting algorithms [15, 16, 17].

We developed and evaluated the GBT and RF voting models using the following procedure. First, we trained the GBT and RF voting models on the *k* = 1, 2, …, 39 best-performing teams on the training set. Next, we chose the value of *k* that achieved the best performance on the validation set; this step identified *k* = 14 for the GBT models and *k* = 2 for the RF models on the murmur detection task and *k* = 14 for the GBT models and *k* = 4 for the RF models on the clinical outcome identification task. Finally, we evaluated the resulting models on the test set.

The GBT and RF voting models performed slightly better than the highest-ranked entry for the murmur detection task (weighted accuracy metric of 0.790 and 0.789, respectively vs. 0.780; see Table 5) and performed slightly worse than the highest-ranked entries for the clinical outcome identification task (cost of 11357 and 11687, respectively vs. 11144; see Table 6). In each case, the voting algorithms had comparable performance to the highest ranked individual entry, but they did not significantly outperform them in either task by any the traditional or novel scoring metrics that we considered.

## Discussion

Cardiac auscultation is one of the most cost-effective tools for helping clinicians to identify heart murmurs, and the CirCor Digiscope dataset enriches our understanding cardiac auscultation within resource-constrained environment. However, despite the novelty and value of this dataset, it, like every dataset, has limitations.

No ages were available for the pregnant individuals in the CirCor DigiScope dataset. It was unclear to the teams if the pregnant individuals belonged to the pediatric age group of the rest of the patients, or if they had a different set of exclusion criteria from the other patients, potentially limiting the performance and appropriateness of models that use this dataset on pregnant individuals.

There was a single human annotator for labeling the heart murmurs; we did not have information about the number of human annotators for labeling the clinical outcomes of abnormal or normal heart function. We expect disagreements between annotators, and several annotators are often needed to produce a single, consistent annotation of a health record. Even several annotators may produce disparate annotations [58]. It is likely that different or more annotators would result in different annotations.

Some heart murmurs are pathological, indicating a physiological problem that requires monitoring and/or intervention. Other heart murmurs are innocent. The human annotator did not identify which heart murmurs were pathological or innocent; the full examination is the only evidence in the dataset that helps to characterize the severity of any of the heart murmurs. We only evaluated the ability of algorithms to identify heart murmurs, and not their ability to identify pathological heart murmurs, but such as task is still valuable for screening.

The definition of a task and the choice of evaluation metrics for quantifying an algorithm’s performance for the task affects the actual and perceived clinical relevance of the algorithms [14]. The definitions of the Challenge evaluation metrics (1) and (8) are attempts to make the evaluation metrics, and therefore the algorithms developed for these metrics, more clinically relevant. The correlation in performance between the traditional and Challenge metrics demonstrate that methods that perform better by one metric tended to perform better by another (at least for the murmur detection task), but the best-scoring method by one metric was often different from the best best-scoring method by another metric, motivating the careful consideration of metrics; see Fig 6 and Fig 7. We also recognize that these metrics are imperfect descriptions of clinical needs; in particular, healthcare cost can be a poor proxy for health needs [13].

The relatively poor performance of the methods on the clinical outcome identification task by all metrics suggests the difficulty of performing this task using the PCG recordings alone; we note that echocardiography is a standard diagnostic tool for assessing cardiac function, which is a more expensive and less accessible modality than phonocardiography.

The voting algorithms had only modest performance improvements, if any, over the individual algorithms. Indeed, the voting algorithms did not use any data-derived features, which could help to provide context by associating different algorithms with different patient subgroups. The features for the voting algorithms included the top *k* algorithms, even when including a lower-ranked entry with a worse overall score or excluding a higher-ranked entry with a better overall score would improve the performance of the voting algorithm. This experiment only considered GBT and RF models, but other types of models could potentially achieve better performance. The training procedure for the GBT and RF models did not use hyperparameter optimization (beyond the selection of the number *k* of individual algorithms), limiting their performance as well. Also, unlike the Challenge teams, we had access to the test set, but, like the Challenge teams, our formal training procedure did not use it. Despite these limitations, the ability of the voting algorithms to slightly improve on the performance on the top-ranked algorithms for the murmur detection task, but not the clinical outcome identification task, also suggests the differences in difficulty between the two related tasks considered by the Challenge.

## Conclusions

This year’s Challenge explored the potential for algorithmic pre-screening for heart murmurs and abnormal cardiac function in resource-constrained environments. We invited the Challenge participants to design working, open-source algorithms for identifying heart murmurs and clinical outcomes from PCG recordings, resulting in 53 working implementations of different algorithmic approaches for interpreting PCGs. A voting approach to combining the diverse approaches resulted in a superior performance over the individual algorithms for murmur detection but not for clinical outcome identification.

By reducing human screening of patients with normal cardiac function, algorithms can lower healthcare costs and increase the effective screening capacity for patients with abnormal cardiac function. The cost function proposed in this Challenge could be the basis for cost-effective screening that balances both the financial and health burden of correctly or incorrectly classifying patients. However, it will be important to optimize the proposed cost function for a given healthcare system or population, since disease prevalence, financial resources, and healthcare resources can differ enormously in different settings.

## Data Availability

All data used in the study are available at https://physionet.org/content/circor-heart-sound/. All data produced in the study will be contained in the manuscript or at https://physionet.org.

## Acknowledgements

This research is supported by the National Institute of General Medical Sciences (NIGMS) and the National Institute of Biomedical Imaging and Bioengineering (NIBIB) under NIH grant numbers 2R01GM104987-09 and R01EB030362 respectively, the National Center for Advancing Translational Sciences of the National Institutes of Health under Award Number UL1TR002378, as well as the Gordon and Betty Moore Foundation and MathWorks under unrestricted gifts. GC has financial interests in AliveCor, LifeBell AI and Mindchild Medical. GC also holds a board position in LifeBell AI and Mindchild Medical. AE receives financial support through grant PID2021-122727OB-I00 funded by MCIN/AEI/10.13039/501100011033 and “ERDF A way of making Europe” and by the Basque Government under Grant IT1717-22. FR and MC receive financial support by National Funds through the Portuguese funding agency, FCT - Fundação para a Ciência e a Tecnologia, within project UIDB/50014/2020. None of the aforementioned entities influenced the design of the Challenge or provided data for the Challenge. The content of this manuscript is solely the responsibility of the authors and does not necessarily represent the official views of the above entities.

## Additional scoring metrics

We defined additional scoring metrics to allow us to make more direct comparisons between methods and tasks.

In particular, we defined a weighted accuracy metric for the clinical outcome identification task as

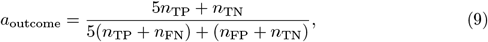

where Table 4 defines a two-by-two confusion matrix *N* = [*n*_*ij*_] for the clinical outcome abnormal and normal classes.

We defined the total cost of diagnosis and treatment with algorithmic pre-screening of murmurs as

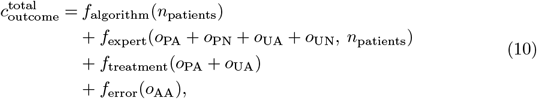

where Table 7 defines a three-by-two confusion matrix *O* = [*o*_*ij*_] for the clinical outcome abnormal and normal classes, *n*_patients_ is the total number of patients, and *f*_algorithm_, *f*_expert_, *f*_treatment_, *f*_error_ are defined above.

**Table 7.**
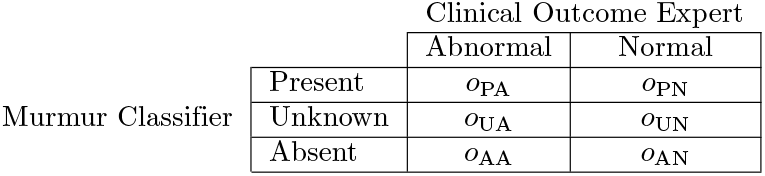
Confusion matrix for murmur detection task with three classes (murmur present, murmur unknown, and murmur absent) using clinical outcomes with two classes (clinical outcome abnormal and clinical outcome normal). The columns are the ground truth labels from the human annotator, and the rows are the classifier outputs. Each entry of the confusion matrix is the number of patients with the classifier outputs for the ground truth labels.

## Mathematical derivation of the cost-based scoring metric

We defined the cost of expert screening to reflect the non-linear costs associated with a limited screening capacity of healthcare systems. While providing fewer screenings incurs a lower total screening cost, the screenings are typically more expensive on a per-screening basis because of the wasted capacity of the system. Similarly, while more screenings incur higher costs, the screenings are also typically more expensive on a per-screening basis because of the inadequate capacity of the system.

Let *s* be the number of expert screenings in a patient cohort of *t* patients, and let *x* = *s/t* be the fraction of the cohort receiving expert screenings. We defined *g*_expert_(*x*) = *a* + *bx* + *cx*^2^ + *dx*^4^ as the mean expert screening cost for screening a fraction *x* of a cohort, and we in turn defined 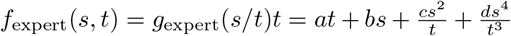 as the total cost for *s* expert screenings in a cohort of *t* patients. These quantity are quartic functions with four unknowns, allowing us to satisfy four criteria:

1. We set *g*_expert_(0) = 25 to define a cost for maintaining the ability to perform expert screening incurs a cost, even when screening *x* = 0 of a cohort, i.e., screening none of the cohort.
2. We set 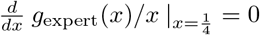 so that mean expert screening cost cost achieved its minimum when screening 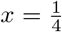 of a cohort, which was roughly half of the prevalence rate of abnormal cases in the database,
3. We set 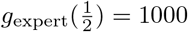 so that the mean expert screening cost was $1000 when screening 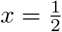 of a cohort, which is roughly the prevalence rate of abnormal cases in the database.
4. We set *g*_expert_(1) = 10000 so that the mean expert screening cost was $10000 when screening *x* = 1, i.e., screening all of the cohort, which is ten times the cost of screening half of the database.

The unique coefficients that satisfy these conditions are *a* = 25, *b* = 397, *c* = −1718, and *d* = 11296.

## Notes

### Clinical Protocols

https://physionetchallenges.org/2022/

### Author Declarations

The study protocol was approved by the 5192-Complexo Hospitalar HUOC/PROCAPE Institutional Review Board, under the request of the Real Hospital Portugues de Beneficencia em Pernambuco.

### Summary of Updates

Results and Discussion substantially updated to reflect the results of the Challenge; various edits made to improve the quality and clarity of the manuscript.

## References

1. Amin DS, Fethi BR. Features for heartbeat sound signal normal and pathological. Recent Patents on Computer Science. 2008;1(1):1–8.

2. Singh J, Anand RS. Computer aided analysis of phonocardiogram. Journal of Medical Engineering & Technology. 2007;31(5):319–323. doi:10.1080/03091900500282772.

3. Marcus G, Vessey J, Jordan MV, Huddleston M, McKeown B, Gerber IL, et al. Relationship Between Accurate Auscultation of a Clinically Useful Third Heart Sound and Level of Experience. Archives of Internal Medicine. 2006;166(6):617–622. doi:10.1001/archinte.166.6.617.

4. Ishmail AA, Wing S, Ferguson J, Hutchinson TA, Magder S, Flegel KM. Interobserver Agreement by Auscultation in the Presence of a Third Heart Sound in Patients with Congestive Heart Failure. Chest. 1987;91(6):870–873. doi:10.1378/chest.91.6.870.

5. Vermarien H. Phonocardiography. In: Webster JG, editor. Encyclopedia of Medical Devices and Instrumentation. vol. 5. 2nd ed. John Wiley & Sons, Ltd; 2006. p. 278–290.

6. Oliveira JH, Renna F, Costa P, Nogueira D, Oliveira C, Ferreira C, et al. The CirCor DigiScope Dataset: From Murmur Detection to Murmur Classification. IEEE Journal of Biomedical and Health Informatics. 2021; p. 1–1. doi:10.1109/jbhi.2021.3137048.

7. Liu C, Springer D, Li Q, Moody B, Juan RA, Chorro FJ, et al. An open access database for the evaluation of heart sound algorithms. Physiological Measurement. 2016;37(12):2181–2213. doi:10.1088/0967-3334/37/12/2181.

8. Oliveira J, Renna F, Mantadelis T, Coimbra M. Adaptive Sojourn Time HSMM for Heart Sound Segmentation. IEEE Journal of Biomedical and Health Informatics. 2019;23(2):642–649. doi:10.1109/jbhi.2018.2841197.

9. Renna F, Oliveira J, Coimbra MT. Deep Convolutional Neural Networks for Heart Sound Segmentation. IEEE Journal of Biomedical and Health Informatics. 2019;23(6):2435–2445. doi:10.1109/jbhi.2019.2894222.

10. Freeman AR. The Clinical Significance of the Systolic Murmur. Annals of Internal Medicine. 1933;6(11):1371. doi:10.7326/0003-4819-6-11-1371.

11. Goldberger AL, Amaral LA, Glass L, Hausdorff JM, Ivanov PC, Mark RG, et al. PhysioBank, PhysioToolkit, and PhysioNet: Components of a New Research Resource for Complex Physiologic Signals. Circulation. 2000;101(23):e215–e220. doi:10.1161/01.CIR.101.23.e215.

12. Reyna MA, Josef C, Jeter R, Shashikumar SP, Westover MB, Nemati S, et al. Early Prediction of Sepsis from Clinical Data: the PhysioNet/Computing in Cardiology Challenge 2019. Critical Care Medicine. 2019;48:210–217. doi:10.1097/CCM.0000000000004145.

13. Mullainathan S, Obermeyer Z. On the Inequity of Predicting A While Hoping for B. In: AEA Papers and Proceedings. vol. 111; 2021. p. 37–42.

14. Reyna MA, Nsoesie EO, Clifford GD. Rethinking Algorithm Performance Metrics for Artificial Intelligence in Diagnostic Medicine. JAMA. 2022;328(4):329–330. doi:10.1001/jama.2022.10561.

15. Friedman JH. Greedy function approximation: a gradient boosting machine. Annals of statistics. 2001; p. 1189–1232.

16. Chen T, Guestrin C. Xgboost: A scalable tree boosting system. In: Proceedings of the 22nd acm sigkdd international conference on knowledge discovery and data mining; 2016. p. 785–794.

17. Breiman L. Random forests. Machine learning. 2001;45:5–32.

18. Lu H, Yip J, Steigleder T, Grießhammer S, Jami NVSJ, Eskofier B, et al. A Lightweight Robust Approach for Automatic Heart Murmurs and Clinical Outcomes Classification from Phonocardiogram Recordings. vol. 49. In Press; 2022. p. 1–4.

19. McDonald A, Gales M, Agarwal A. Detection of Heart Murmurs in Phonocardiograms with Parallel Hidden Semi-Markov Models. vol. 49. In Press; 2022. p. 1–4.

20. Xu Y, Bao X, Lam HK, Kamavuako E. Hierarchical Multi-Scale Convolutional Network for Murmurs Detection on PCG Signals. vol. 49. In Press; 2022. p. 1–4.

21. Krones F, Walker B, Mahdi A, Kiskin I, Lyons T, Parsons G. Dual Bayesian ResNet: A Deep Learning Approach to Heart Murmur Detection. vol. 49. In Press; 2022. p. 1–4.

22. Lee J, Kang T, Kim N, Han S, Won H, Gong W, et al. Deep Learning Based Heart Murmur Detection using Frequency-time Domain Features of Heartbeat Sounds. vol. 49. In Press; 2022. p. 1–4.

23. Alkhodari M, Azman S, Hadjileontiadis L, Khandoker A. Ensemble Transformer-Based Neural Networks Detect Heart Murmur In Phonocardiogram Recordings. vol. 49. In Press; 2022. p. 1–4.

24. Monteiro S, Fred A, Silva H. Detection of Heart Sound Murmurs and Clinical Outcome with Bidirectional Long Short-Term Memory Networks. vol. 49. In Press; 2022. p. 1–4.

25. Ľubomír Antoni, Bruoth E, Szabari A, Vozáriková G, Bugata P, Jr PB, et al. Murmur Identification Using Supervised Constrastive Learning. vol. 49. In Press; 2022. p. 1–4.

26. Nejedly P, Pavlus J, Smisek R, Koscova Z, Vargova E, Viscor I, et al. Classification of heart murmurs using an ensemble of residual CNNs. vol. 49. In Press; 2022. p. 1–4.

27. Summerton S, Wood D, Murphy D, Redfern O, Benatan M, Kaisti M, et al. Two-stage Detection of Murmurs from Phonocardiograms using Deep and One-class Methods. vol. 49. In Press; 2022. p. 1–4.

28. Bai Z, Yan B, Chen X, Wu Y, Wang P. Murmur Detection and Clinical Outcome Classification Using a VGG-like Network and Combined Time-Frequency Representations of PCG Signals. vol. 49. In Press; 2022. p. 1–4.

29. Rohr M, Müller B, Dill S, Güney G, Antink CH. Two-Stage Multitask-Learner for PCG Murmur Location Detection. vol. 49. In Press; 2022. p. 1–4.

30. Ballas A, Papapanagiotou V, Delopoulis A, Diou C. Listen to your heart: A self-supervised approach for detecting murmur in heart-beat sounds. vol. 49. In Press; 2022. p. 1–4.

31. Wen H, Kang J. Searching for Effective Neural Network Architectures for Heart Murmur Detection from Phonocardiogram. vol. 49. In Press; 2022. p. 1–4.

32. Costa J, Rodrigues R, Couto P. Multitask and Transfer Learning for Murmur Detection in Heart Sounds. vol. 49. In Press; 2022. p. 1–4.

33. Knorr M, Bremer J, Schnabel R. Using Mel-Spectrograms and 2D-CNNs to detect Murmurs in Variable Length Phonocardiograms. vol. 49. In Press; 2022. p. 1–4.

34. Araujo M, Zeng D, Palotti J, Hu X, Shi Y, Pyles L, et al. Maiby’s Algorithm: A Two-stage Deep Learning approach for Murmur Detection in Mel Spectrograms for Automatic Auscultation of Congenital Heart Disease. vol. 49. In Press; 2022. p. 1–4.

35. Xia P, Yao Y, Liu C, Zhang H, Wang Y, Xu L, et al. Heart Murmur Detection from Phonocardiogram Based on Residual Neural Network with Classes Distinguished Focal Loss. vol. 49. In Press; 2022. p. 1–4.

36. Venkataramani VV, Garg A, Priyakumar UD. Modified Variable Kernel Length ResNets for Heart Murmur Detection and Clinical Outcome Prediction using Multi-positional Phonocardiogram Recording. vol. 49. In Press; 2022. p. 1–4.

37. Chang Y, Liu L, Antonescu C. Multi-Task Prediction of Murmur and Outcome from Heart Sound Recordings. vol. 49. In Press; 2022. p. 1–4.

38. Kim J, Park G, Suh B. Classification of Phonocardiogram Recordings using Vision Transformer Architecture. vol. 49. In Press; 2022. p. 1–4.

39. Shin JM, Kim HS, Seo WY, Kim SH. Learning Time-Frequency Representations of Phonocardiogram for Murmur Detection. vol. 49. In Press; 2022. p. 1–4.

40. Peng G, Zou H, Wang J. Classification of phonocardiograms using residual convolutional neural network and MLP. vol. 49. In Press; 2022. p. 1–4.

41. Petrolis R, Paukstaitiene R, Rudokaite G, Macas A, Grigaliunas A, Krisciukaitis A. Convolutional neural network aproach for heart MurMur detection in auscultation signals using wavelet transform based features. vol. 49. In Press; 2022. p. 1–4.

42. Gemke P, Spicher N, Kacprowski T. An LSTM-based Listener for Early Detection of Heart Disease. vol. 49. In Press; 2022. p. 1–4.

43. Hu L, Cai W, Li X, Li J. Detection of murmurs from heart sound recordings with deep residual networks. vol. 49. In Press; 2022. p. 1–4.

44. Warrick P, Afilalo J. Phonocardiographic Murmur Detection by Scattering-Recurrent Networks. vol. 49. In Press; 2022. p. 1–4.

45. Imran Z, Grooby E, Sitaula C, Malgi V, Aryal S, Marzbanrad F. A Fusion of Handcrafted Features and Deep Learning Classifiers for Heart Murmur Detection. vol. 49. In Press; 2022. p. 1–4.

46. Wang X, Fan F, Gao H, Zhang S, Yang C, Li J, et al. Beat-wise uncertainty learning for murmur detection in heart sounds. vol. 49. In Press; 2022. p. 1–4.

47. Singstad BJ, Bongo L, Johnsen M, Ravn J, Gitau A, Schirmer H. Phonocardiogram Classification Using 1-Dimensional Inception Time Convolutional Neural Networks. vol. 49. In Press; 2022. p. 1–4.

48. Choi S, Seo HC, Choi K, Yoon G, Joo S. Murmur Classification with U-net State Prediction. vol. 49. In Press; 2022. p. 1–4.

49. Li X, Schlindwein F, Ng GA. Transfer Learning in Heart Sound Classification using Mel spectrogram. vol. 49. In Press; 2022. p. 1–4.

50. Cornely A, Mirsky G. Heart Murmur Detection Using Wavelet Time Scattering and Support Vector Machines. vol. 49. In Press; 2022. p. 1–4.

51. Gao Y, Qiao L, Li Z. Heart Murmur Detection of PCG Using ResNet with Selective Kernel Convolution. vol. 49. In Press; 2022. p. 1–4.

52. Ding J, Li JJ, Xu M. Classification of Murmurs in PCG Using Combined Frequency Domain and Physician Inspired Features. vol. 49. In Press; 2022. p. 1–4.

53. Jalali K, Saket MA, Noorzadeh S. Heart Murmur Detection and Clinical Outcome Prediction using Multilayer Perceptron Classifier. vol. 49. In Press; 2022. p. 1–4.

54. Stein G, Festag S, Büchner T, Shadaydeh M, Denzler J, Spreckelsen C. Outcome Prediction and Murmur Detection in Sets of Phonocardiograms by a Deep Learning-Based Ensemble Approach. vol. 49. In Press; 2022. p. 1–4.

55. Rudman W, Merullo J, Mercurio L, Eickhoff C. ACQuA: Anomaly Classification with Quasi-Attractors. vol. 49. In Press; 2022. p. 1–4.

56. Fan P, Shu Y, Han Y. Transformer embedded with learnable filters for heart murmur detection. vol. 49. In Press; 2022. p. 1–4.

57. Bondareva E, Han J, Xia T, Mascolo C. Towards Uncertainty-Aware Murmur Detection in Heart Sounds via Tandem Learning. vol. 49. In Press; 2022. p. 1–4.

58. Clifford GD, Liu C, Moody B, Lehman LwH, Silva I, Li Q, et al. AF Classification from a Short Single Lead ECG Recording: the PhysioNet/Computing in Cardiology Challenge 2017. In: 2017 Computing in Cardiology (CinC). vol. 44. IEEE; 2017. p. 1–4.

